# Empirical transmission advantage of the D614G mutant strain of SARS-CoV-2

**DOI:** 10.1101/2020.09.22.20199810

**Authors:** Kathy Leung, Yao Pei, Gabriel M Leung, Tommy TY Lam, Joseph T Wu

**Affiliations:** WHO Collaborating Centre for Infectious Disease Epidemiology and Control, School of Public Health, LKS Faculty of Medicine, The University of Hong Kong, Hong Kong SAR, China; State Key Laboratory of Emerging Infectious Diseases, School of Public Health, The University of Hong Kong, Hong Kong SAR, China; Joint Institute of Virology (Shantou University and The University of Hong Kong), Guangdong-Hongkong Joint Laboratory of Emerging Infectious Diseases, Shantou University, Shantou, China

## Abstract

The SARS-CoV-2 lineage carrying the amino acid change D614G has become the dominant variant in the global COVID-19 pandemic. The rapid spread of the G614 mutant suggests that it may have a transmission advantage over the D614 wildtype. Using our previous epidemiological framework to analyze COVID-19 surveillance and sequence data, we estimated that the G614 mutant is 31% (28-34%) more transmissible than the D614 wildtype. As such, interventions that were previously effective in containing or mitigating the D614 wildtype (e.g. in China, Vietnam, Thailand, etc.) might be less effective against the G614 mutant. Our framework can be readily integrated into current COVID-19 surveillance to monitor the emergence and fitness of mutant strains, such that pandemic surveillance, disease control and development of treatment and vaccines can be adjusted dynamically.

---

Recent studies of SARS-CoV-2 genomes have identified various mutations associated with different emerging genetic clades. Two major clades were initially reported near the end of first wave of COVID-19 outbreak in China ^1^, and soon the development into pandemic was accompanied by reports of several more clades featuring different mutations among the countries ^2^. Some clades are found to be associated with difference in viral phenotypes and immunological reaction from patients ^3^, highlighting the viral genetic determinants of the outbreak progression and management, and importance of monitoring and assessing emerging variants of SARS-CoV-2.

One of the notable variations, the D614G mutation, encodes a change from aspartic acid to glycine in the carboxy-terminal region of the S1 domain of the viral spike protein of SARS-CoV-2. Notably, the detection of the mutant G614 has increased rapidly since late February 2020 and G614 is now the dominant subtype circulating in most parts of world ^4-7^. The rapid spread of G614 suggests it may have a transmission advantage over the wildtype D614 in terms of faster growth rate due to higher reproductive number or shorter generation time or both ^8^. This hypothesis is corroborated by several *in vitro* studies which showed that the D614G mutation is correlated with increased infectivity in cell models ^9-12^. Recent phylogenetic analysis also showed the evidence of diversifying selection at the site 614, suggesting the potential transmission advantage of D614G ^13,14^. However, limited assessment has been conducted to date to quantify the epidemiological fitness of G614 compared with its wildtype predecessor D614 ^15^. Here we used our previous epidemiological framework for fitness inference of influenza strains ^16^ to analyze COVID-19 surveillance and sequence data and characterize the comparative transmissibility of the G614 mutant.

## Results

### Identification of D614 and G614 co-circulating clusters

The global phylogeny of SARS-CoV-2 shows multiple genetic clades and their associated genomic mutations, of which the clade with G614 mutation is by far the largest (Figure 1). We assumed that the mutation D614G is the only site of interest that potentially confers a transmission advantage and obtained 35,377 sequences collected between 24 December 2019 and 8 June 2020 which covered the 614th position (i.e. either D614 or G614) in the translated amino acid sequences of the spike gene ^4^. We identified phylogenetic clusters of local transmissions in each country from the context of global SARS-CoV-2 phylogeny (see Methods and Figure S1). Each cluster approximately stemmed from one or a small number of introduction events, and included at least two sequences by our definitions. We included countries with both D614 and G614 variants co-circulated in their respective (phylogenetically evidenced) locally sustained transmission clusters for a period of at least two weeks (i.e. at least two disease generations, assuming the mean generation time of 5-7 days). To minimize potential bias due to stochasticity in sampling, we only included countries with 100 or more sequences during the co-circulation period. Ten countries, namely Australia, Belgium, Denmark, Iceland, India, Netherlands, Spain, Portugal, the UK, and the US fulfilled these criteria and were included in our further analysis. In these ten countries, 515 D614 clusters and 1,420 G614 clusters among 10,915 sequences were identified, and the G614:D614 ratio increased over time and the G614 mutant rapidly became the dominant strain in these countries (Figure 2).

**Figure 1.**
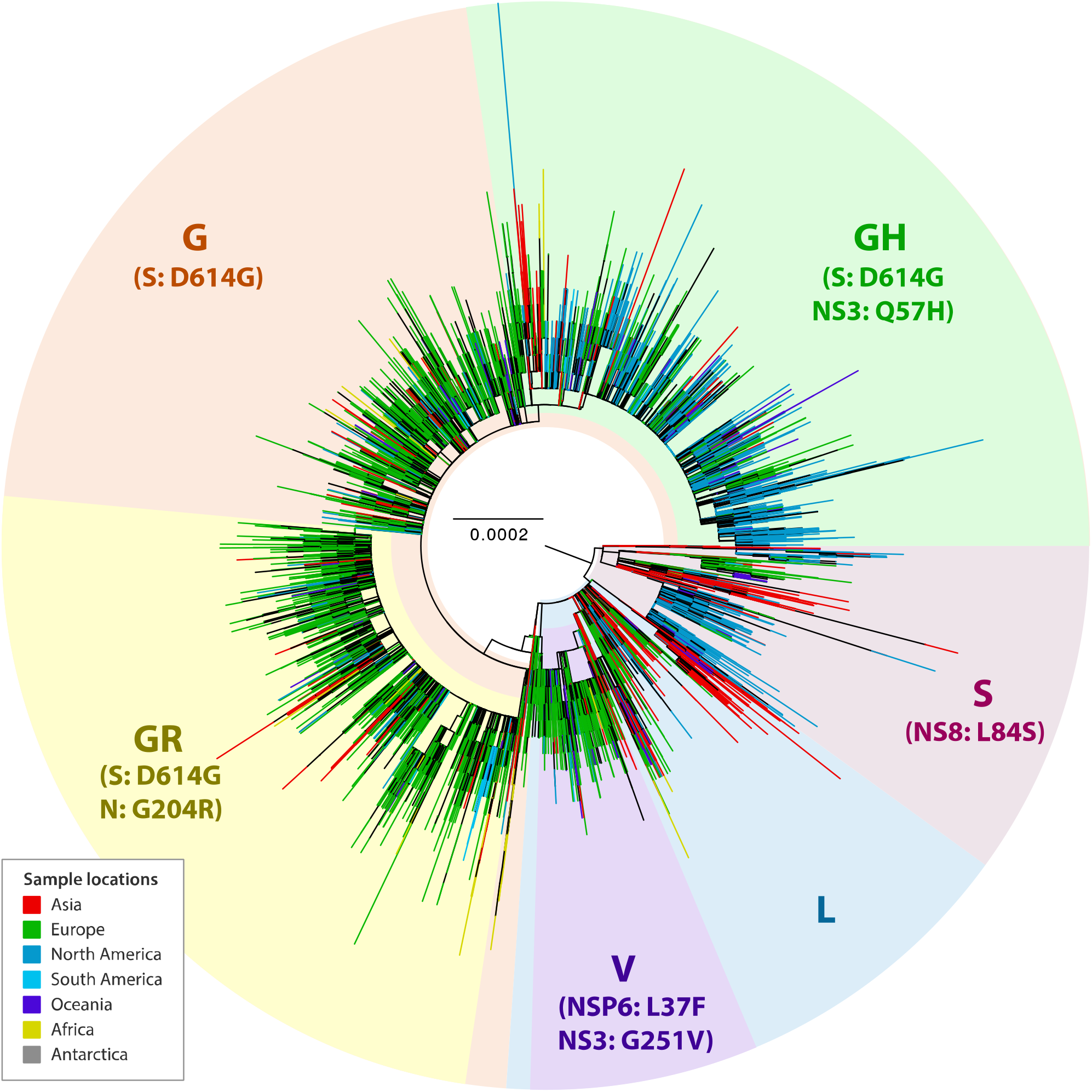
Global phylogeny of SARS-CoV-2. The maximum likelihood tree was inferred from the alignment of 26,244 worldwide SARS-CoV-2 genome sequences with high sequencing coverage, using GTR+CAT substitution model in FastTree program. Multiple clades are highlighted, and their associated mutations are indicated within parenthesis. Tree tips corresponding to the viral sequences from different continents are annotated in different colours as shown in the colour legend box.

**Figure 2.**
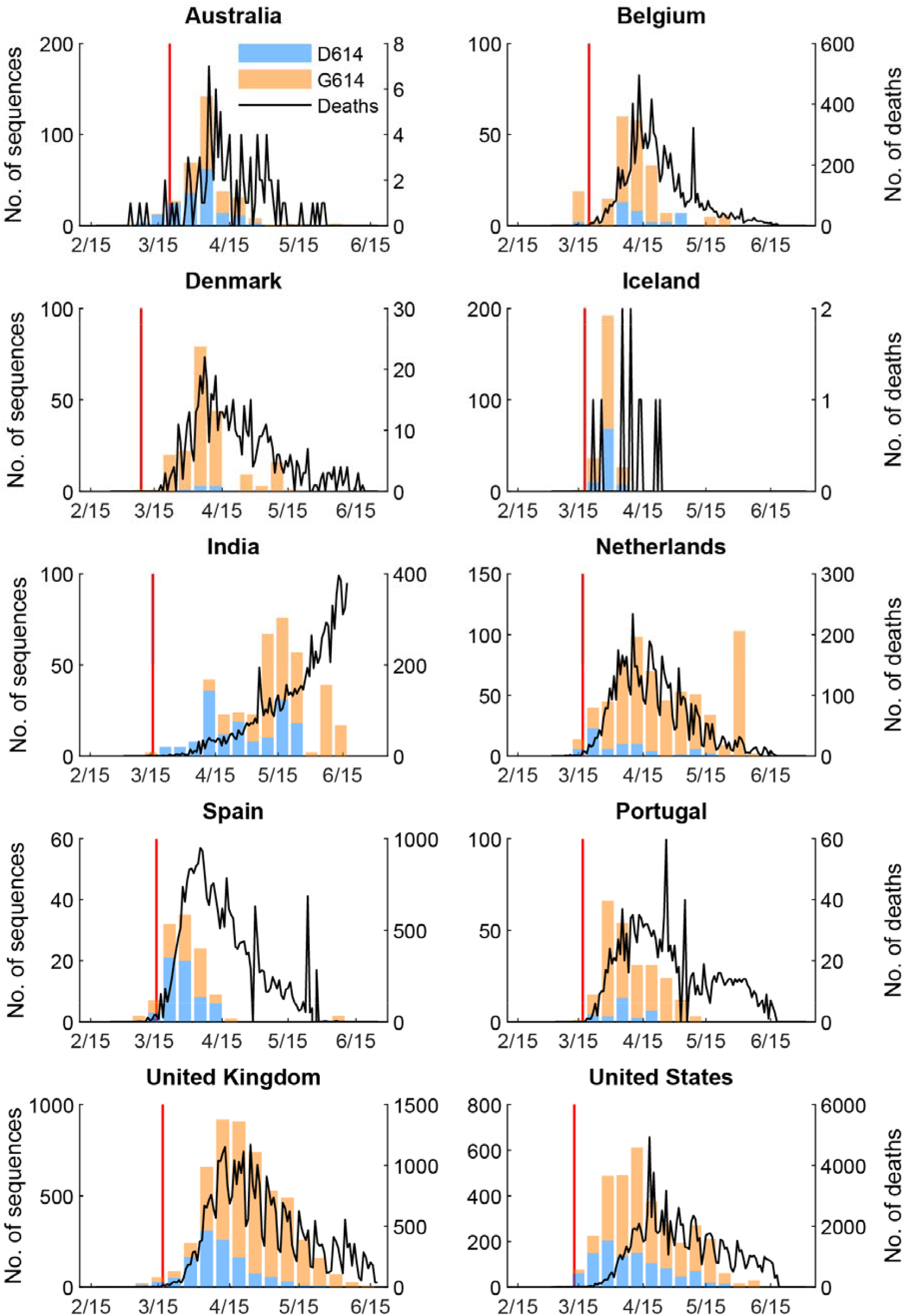
Weekly number of confirmed COVID-19 deaths and the weekly number of D614 and G614 sequences from phylogenetically defined transmission clusters, submitted by Australia, Belgium, Denmark, Iceland, India, Netherlands, Spain, Portugal, UK and US. Clusters with 2 or more sequences on GISAID were defined using phylogenetic methods with “strict” criteria (See Methods and Figure S1). Each cluster stemmed from one or a small number of introductions and at least one transmission chain can be reconstructed from sequences within the same cluster. Only clusters sampled during the co-circulating period of D614 and G614 for at least two weeks in each country were included in the analysis. Only countries with more than 100 sequences were included in the analysis. The first/index case of each cluster was included in the analysis. The red lines indicated the date when major travel restriction from or to countries of European Union started.

### Inference of the G614 fitness in transmission

Let *σ* be the ratio of the basic reproduction number of the G614 strain to that of the D614 strain, and the ratio of the mean generation time of the G614 strain to that of the D614 strain. We assumed that the mean generation time of the D614 strain was 5.4 days ^17^. Given that the G614 mutant has displaced the D614 wildtype globally, we assumed *σ*≥1 and *τ*≤1. Using confirmed deaths (adjusted for the delay between onset and death) as the proxy for the COVID-19 epidemic curve, we estimated that was 1.31 (95% CrI 1.28-1.34) and *τ*was 0.99 (0.96-1.00) across the ten countries. That is, the basic reproductive number of the G614 mutant was 31% (28-34%) higher than that of the D614 wildtype, and the mean generation time of the two strains were essentially the same. The fitted model was congruent with the observed proportions of G614 isolates over time in all ten countries (Figure 3). If we used confirmed cases instead of confirmed deaths as the proxy for the COVID-19 epidemic curve (Figure S2 and Figure S3), then *σ* was 1.23 (95% CrI 1.19-1.26) and *τ* was 0.96 (0.90-1.00).

**Figure 3.**
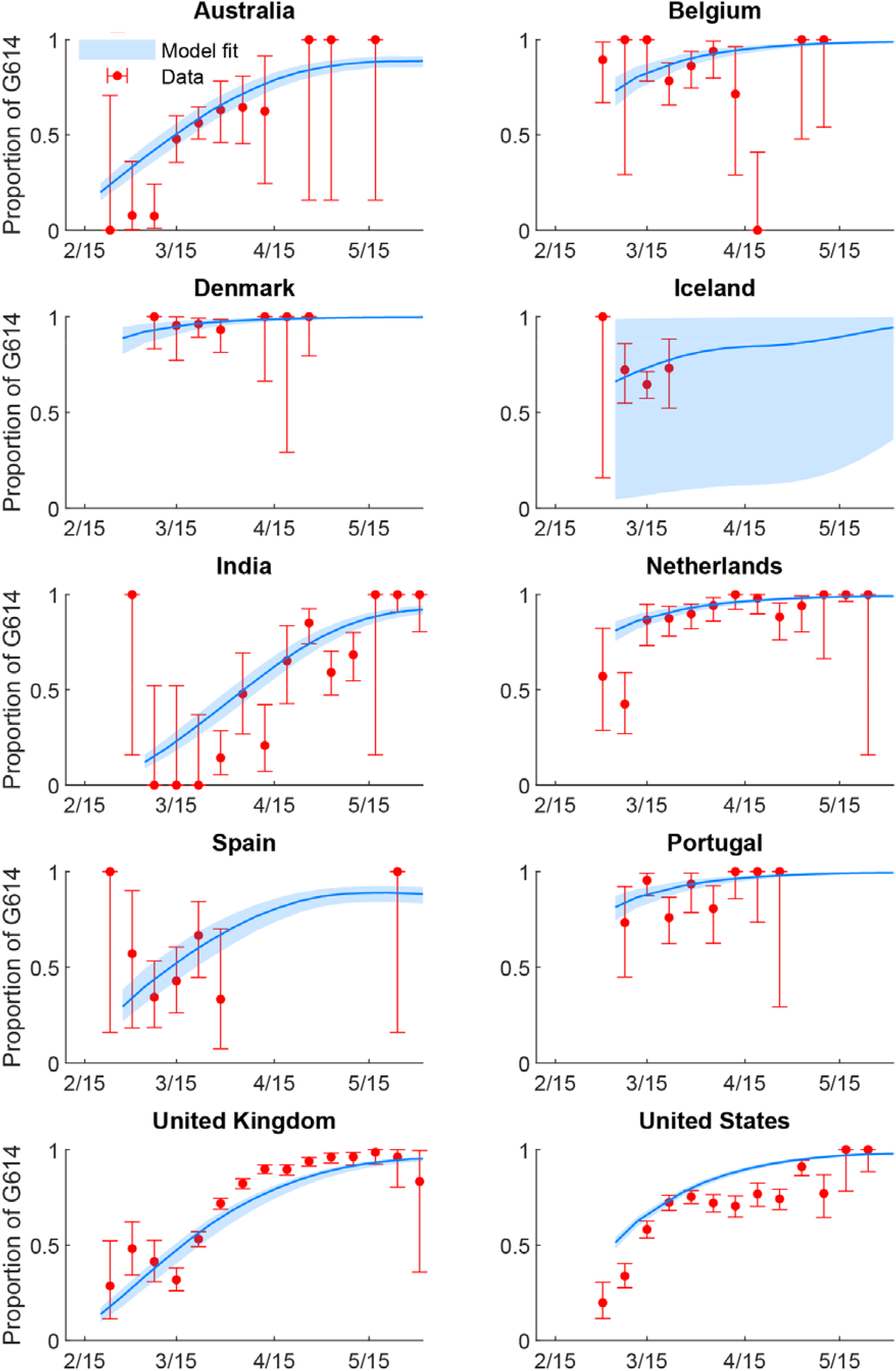
The weekly proportion of G614 sequences between late January and early May when both D614 and G614 strains cocirculated. The time series of confirmed COVID-19 deaths was used in the estimation. The red circles and error bars indicated the observed proportion with 95% binomial CIs among sequence data. The blue lines and shades indicated the posterior mean and 95% CrI of the estimates.

To assess potential geographical heterogeneity in the transmission advantage of the G614 mutant, we allowed *σ* to differ among the US, the UK and the remaining locations and reran the inference. The resulting estimates for *σ* was 1.13 (1.09-1.16), 1.53 (1.28-1.58), and 1.30 (1.19-1.42) for the US, the UK,and other locations, respectively, with *τ* = 0.99 (0.93-1.00).

In the fitness estimation, by reconstructing co-circulating clusters of D614 and G614 using phylogenetic methods, we were able to exclude sequences from importations that generated no or limited secondary infections. However, the global phylogeny of SARS-CoV-2 suggested that most countries in Europe (such as UK ^18^) and the US received overwhelming importations of G614 since late February, possibly from countries with largely undetected outbreaks dominated by G614. To assess the effects of dominant introductions of G614, we incorporated G614 importation in the fitness estimation by specifically assuming the imported infections consisted of G614 only and the imported G614 force of infection was *φ*^*G*^ times of the local COVID-19 incidence rate (see Methods). We performed a sensitivity analysis on sequences from the UK, which is among the countries with the largest number of SARS-CoV-2 genomes made available to the public (Figure 4). The resulting estimate of *φ* _*G*_ was 0.0012 (0.001-0.0035), suggesting that the dominant G614 importations was not driving the increase of G614 over time in UK (Figure S4). Similarly, assuming *φ* _*G*_ was the same in all the ten selected countries, the resulting *φ*_*G*_ estimate was 0.0172 (0.0028-0.0271, Table S1).

**Figure 4.**
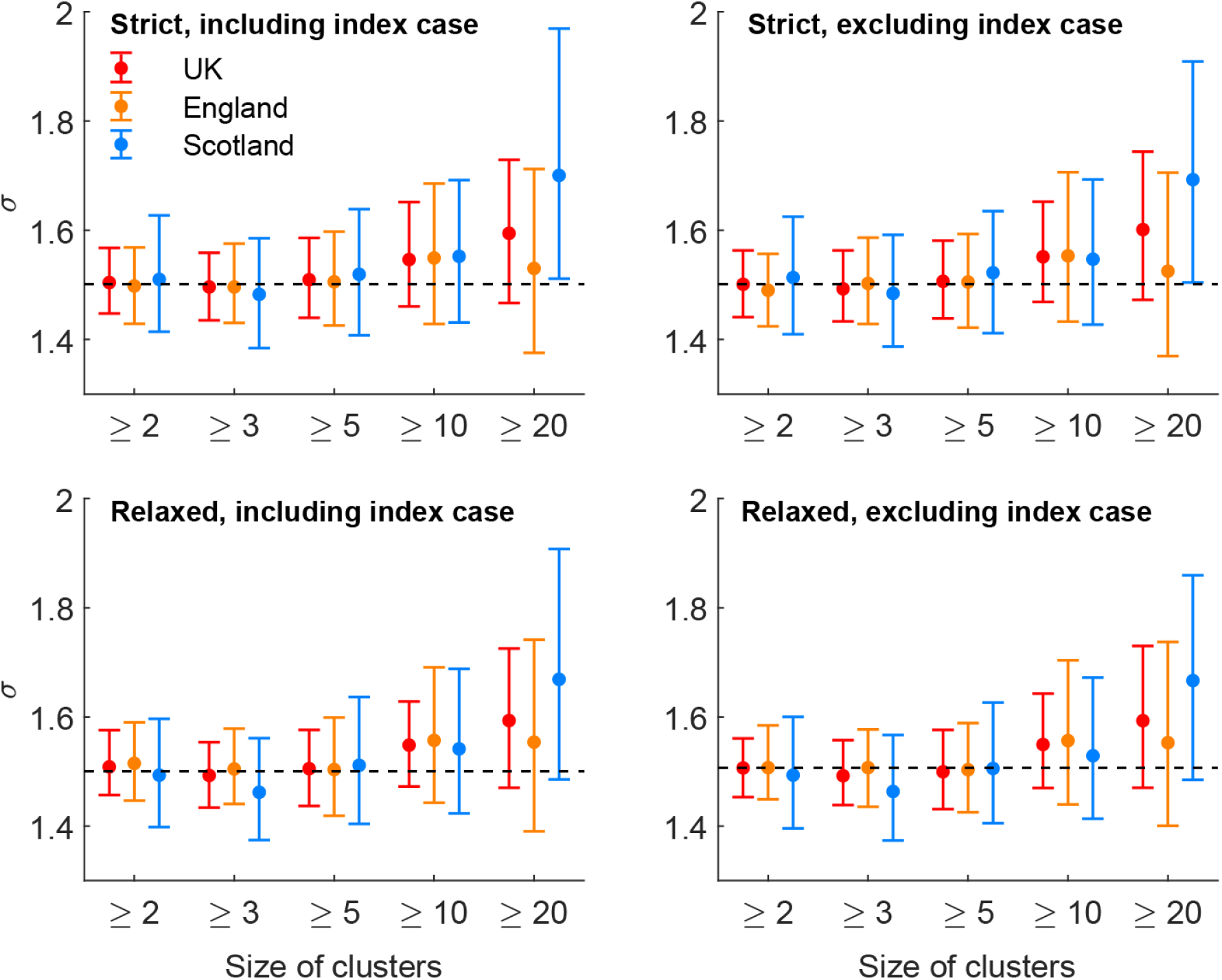
Estimates of G614 fitness in England and Scotland under different phylogenetic definitions and minimum sizes of local transmission clusters. (A) Base case as in Figure 2 and 3, including clusters with ≥2, ≥3, ≥5, ≥10 and ≥20 sequences in each cluster using the “strict” cluster definition assuming index case in each cluster was included. (B) Including clusters with ≥2, ≥3, ≥5, ≥10 and ≥20 sequences in each cluster using the “strict” cluster definition assuming index case in each cluster was excluded. (C) Including clusters with ≥2, ≥3, ≥5, ≥10 and ≥20 sequences in each cluster using the “relaxed” cluster definition assuming index case in each cluster was included. (D) Including clusters with ≥2, ≥3, ≥5, ≥10 and ≥20 sequences in each cluster using the “relaxed” cluster definition assuming index case in each cluster was excluded. The time series of confirmed COVID-19 deaths was used in the estimation. The circles and error bars indicated the posterior mean and 95% CrI of G614 fitness estimates. The horizontal dashed line showed the posterior mean of G614 fitness estimates of UK in the base case (including clusters with ≥2 sequences in each cluster using the “strict” cluster definition assuming index case in each cluster was included; including clusters reconstructed from England, Scotland, Wales and Northern Ireland).

Although G614 introductions occurred later, more clusters with G614 were reconstructed in the ten selected countries and these clusters were larger on average. However, the size of clusters strongly depended on the sampling scheme and sequencing priority in each country. To assess the effects of sampling frequency in the G614 fitness estimation, we performed a sensitivity analysis on sequences from the UK. We included only clusters with at least 2, 3, 5, 10 or 20 different patient sequences in the fitness estimation (Figure 4). We found that estimates of were not sensitive to the minimum cluster sizes up to 20 sequences. The estimations of were also not sensitive to the definitions of phylogenetic topology (i.e. “strict” and “relaxed” definitions; see Methods and Figure S1) used to identify the D614 and G614 local transmission clusters (Figure 4).

### Effects of G614 fitness in the SARS-CoV-2 transmission dynamics

The inferred value of *σ* suggests that the herd immunity threshold for the G614 mutant is higher than that for the D614 wildtype. For example, if mixing is homogeneous, the excess is 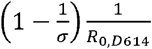 where R_0,D614_ is the basic reproductive number of the D614 wildtype. Using the inferred value of *σ* =1.31, we estimated that the D614G mutation would increase the herd immunity threshold from 50% to 62% (i.e. 12% excess) if *R*_0,*D*614 *=*_2 and from 67% to 75% (i.e. 8% excess) if *R*_*0,D614 =*_3. More robust estimates of herd immunity threshold would require accounting for heterogeneities in age-dependent physical mixing, susceptibility, infectiousness, etc. ^19^

Although the above results suggested that there is no difference between the generation time of the two strains, we conducted a sensitivity analysis to assess the possibility that the transmission advantage oG614 was entirely due to shorter generation time, i.e. *τ*<1 and *σ*=1. The resulting estimate of *τ* was 0.80 (0.75-0.86), i.e. the mean generation time of G614 was 20% (14-25%) shorter than that of D614. However, this fitted model had significantly higher AIC than our base case model, hence supporting our base case conclusion that the mean generation time of the two strains were essentially the same and the transmission advantage of the G614 mutant was entirely due to higher infectivity.

Compared with Australia and US, the countries in Europe suffered from earlier introduction of the G614 strain (Table 1). The proportion of G614 infections reached 19-74% in late February to early March for countries in Europe. Similarly, more detailed breakdown of US data showed that New York State had earlier introduction of G614 compared with Washington State. Assuming *τ*=1, we estimated that was 1.25 (1.20-1.30) for the Washington State, but the G614 fitness was not estimated for the New York State due to the lack of co-circulating clusters of both strains.

**Table 1.** The proportion of G614 infections when both D614 and G614 started to cocirculate.

| **Country** | **GISAID ID of the 1<sup>st</sup> sequence in D614 and G614 cocirculating clusters included in the analysis** | **Sampling date of the 1<sup>st</sup> sequence in D614 and G614 cocirculating clusters included in the analysis** | **$\rho(0)$ (95% CrI)** |
| --- | --- | --- | --- |
| Australia | EPI_ISL_420456 | 22 February 2020 | 0.132 (0.10-0.169) |
| Belgium | EPI_ISL_415155 | 1 March 2020 | 0.622 (0.528-0.714) |
| Denmark | EPI_ISL_416143 | 28 February 2020 | 0.834 (0.720-0.919) |
| Iceland | EPI_ISL_427757 | 6 March 2020 | 0.501 (0.023-0.975) |
| India | EPI_ISL_420543 | 3 March 2020 | 0.071 (0.050-0.098) |
| Netherlands | EPI_ISL_413588 | 1 March 2020 | 0.735 (0.665-0.798) |
| Spain | EPI_ISL_418251 | 25 February 2020 | 0.192 (0.135-0.264) |
| Portugal | EPI_ISL_418011 | 4 March 2020 | 0.738 (0.649-0.816) |
| United Kingdom | EPI_ISL_466615 | 16 February 2020 | 0.071 (0.048-0.096) |
| United States | EPI_ISL_417100 | 29 February 2020 | 0.384 (0.349-0.417) |

## Discussion

Our findings suggest that SARS-CoV-2 strain with the G614 mutation is 31% more transmissible than the wildtype D614 strain. Such increase in fitness propels the G614 strain to displace the wildtype D614 strain and became the dominant strain in Europe within 2 months after its first detection. Our findings are consistent with the differential growth rates of D614 and G614 lineages estimated from the a different phylodynamic analysis in UK ^7,15^: G614 lineages grow at a rate 1.21 (logistic model; 95% CI 1.06-1.56) times of that of D614. Our results are also largely consistent with the rate at which COVID-19 was resurging in Beijing in comparison to the spread of the D614-dominant first wave in January-February. Whole genome sequencing showed that the strain causing the June wave in Beijing was genetically closest to the virus isolates in Europe with G614 ^20^. While 156 local cases were reported between 12 and 31 January for the D614-dominant first wave, 325 local cases were reported between 11 and 30 June for the G614-dominant outbreak. This suggests that the latter was more transmissible given that Beijing had remained extremely vigilant with COVID-19 surveillance and control since mid-January, though intensive community testing was organized only in June and thus more mild infections might have been identified.

We estimated that IFRs were not statistically significant in locations where COVID-19 circulation was predominated by G614, though data were limited (Table S2). Although the virus with G614 seems to cause more mild and asymptomatic infections in Beijing’s Xinfadi outbreak, intensive community testing was organized only in June (and thus more mild infections might have been identified) ^21^: 96.1% (246/256) of confirmed cases were mild or moderate in June, which was higher than 86.7% (216/249) during the first wave ^22^; 7.9% (22/278) of confirmed infections were asymptomatic in June compared with 5.0% (13/262) during first wave ^22^.

Our base case results suggest that *R*_0_ of the G614 strain would be approximately 1.3 times that of the D614 strain which had been estimated to be 2-2.5 using Wuhan data ^23,24^. This is consistent with the recent *R*_*0*_ estimates of 3-4.5 in Europe and US where G614 is dominant^25,26^. Taken together, these results imply that control measures that were sufficient for controlling D614-dominant outbreaks in mainland China would only be 70% as effective against G614-dominant outbreaks. For instance, social distancing interventions were reported to reduce 79% of contacts in Shanghai during the first wave ^27^, which might not be sufficient for Shanghai’s fast and successful suppression of the first wave by mid-February if *R*_0_ were 3-4.5. By the same token, the critical vaccination coverage (which is equivalent to the herd immunity threshold) for G614 would be higher than that for D614. An alternative and less probable explanation for the faster doubling time of the G614 strain was that there was no change in *R*_0_ but the mean generation time of the G614 mutant was around 20% shorter than that of the D614 wildtype. Using the first wave data of mainland Chinese city Guangzhou, we previously estimated that possibly 44% of all COVID-19 infection events were pre-symptomatic transmission and 95% of all transmission would have taken place by day 5 after symptom onset ^28^. If the G614 virus were to spread faster but cause slightly milder illness, its current dominance would require more rapid response (20% faster) in contact tracing and testing to control any outbreak even at the very early stage. However, in this scenario, the critical vaccination coverage for the two strains would be the same because there is no difference in *R*_0_ ^29^.

Our study has several limitations. First, we only considered the D614G mutation and simply categorized the sequences on GISAID by aligning the spike protein region that contains the locus. We did not consider mutations in other loci that might provide necessary genetic background for D614G and act synergistically to affect the fitness of G614. The mutant D614G was detected sporadically among local cases in mainland Chinese provinces Guangdong and Zhejiang after February, but no sustained circulation of G614 clusters had been detected in mainland China until the recent Xinfadi outbreak in Beijing in June. The biological mechanism of increased spread of G614 is still unclear. Second, we estimated the date of infection approximately by deconvoluting the time series of the dates of sampling for sequence data or the dates of reporting of confirmed cases or deaths. Given the relatively high fitness advantage of G614, the date of exposure or symptom onset should be used instead of the date of sampling to generate more accurate fitness estimates, if clinical data of patients could be linked with sequences available on GISAID. Third, our fitness estimation is only applicable when D614 and G614 strain cocirculates, and therefore cannot be used to monitor the fitness of a newly emerged mutant strain that has not yet spread in the community or has already dominated the community transmission. Fourth, our method compares the relative fitness of two strains. We did not consider the scenario while three or more strains cocirculate and their transmissions might interfere with each other. Although sustained G614 transmission was not detected previously in Guangdong and Zhejiang, the mutant strain might have accumulated several necessary mutations chronologically and exhibited a gradual increase in fitness over time. Categorizing all the sequences by D614 and G614 might have oversimplified the biological process and mechanism.

In conclusion, we have shown that the G614 mutation confers a transmission advantage over the wildtype D614. Monitoring the emergence of mutations and fitness of mutant strains are essential during the COVID-19 pandemic because the spread of mutants can attenuate the effectiveness of outbreak response and control interventions, such as development of therapy and vaccines. It is also important to acquire thorough understanding of viral phenotypes, clinical and epidemiological characteristics of emerging mutants like D614G of SARS-CoV-2, such that surveillance and disease control measures could be adjusted dynamically to counter the evolving risks posed by dominant mutant clades. Our method can be readily integrated into the analysis of phylogenetic data in the current COVID-19 surveillance system, to provide efficient epidemiological assessment of the transmission potential of emerging mutants for early alert.

## Methods

### Phylogenetic reconstruction of D614 and G614 clusters

For the convenience of mutation analysis, we first downloaded all the SARS-CoV-2 sequences submitted on or before 15 June 2020 from GISAID ^2^. Multiple sequence alignment was constructed from the downloaded sequences. Then we labeled each sequence with either “D614” or “G614” based on the amino acid found at the 614th position in the translated amino acid sequences of the spike gene ^4^. We excluded sequences with no explicit sample collection dates. In total, 35,377 sequences collected between 24 December 2019 and 8 June 2020 were used to construct the dataset. A phylogenetic tree was built from these global sequences with high sequencing coverages of the genomes, using maximum likelihood heuristic search and GTR+CAT nucleotide substitution model in FastTree v2.1.11 ^30^.

We examined the global phylogeny to identify the different local transmission chains of D614 and G614 in each country, for the use in the fitness model described below. A strict monophyletic lineage of virus strains from the same country was defined as a local transmission cluster (hereinafter “strict” definition, Figure S1). A minimum of two sequences in such a cluster was considered as significant local transmission. We included countries with such clusters respectively of D614 and G614 that have co-circulated for a period at least two weeks (i.e. at least two disease generations, assuming the mean generation time of 5-7 days). To avoid potential bias due to stochasticity in sampling, we only included countries with 100 or more sequences during the co-circulation period. We identified 515 D614 clusters and 1,420 G614 clusters among 10,915 sequences in ten selected countries, namely Australia, Belgium, Denmark, Iceland, India, Netherlands, Spain, Portugal, the UK and the US. We also examined the effect of different cutoffs for minimum cluster size (2, 3, 5, 10 and 20) in our inference.

Since the SARS-CoV-2 genomes evolved in a relatively slower rate and were intensively sampled, there were many unresolved polytomic nodes in the phylogeny and identical sequences from different countries ^31^. This could potentially break a larger local transmission cluster into multiple smaller ones based on the above-mentioned “strict” definition. As such, we also considered a “relaxed” definition under which cluster and non-cluster sequences were grouped into an aggregated cluster if they shared the same parent nodes (Figure S1). We evaluated the sensitivity of our fitness estimates to the “strict” and “relaxed” definitions. We also evaluated the sensitivity to the inclusion or exclusion of earliest sequence in each cluster which may represent the potential index case for the cluster and was less likely derived from the local sustained transmission chains.

### The model

We assume that the mutation D614G is the only site of interest that results in potential difference in transmission advantage of SARS-CoV-2 throughout our analysis. We define the fitness of G614 as the ratio of the basic reproduction number of the strain with G614 to the strain with D614, i.e. 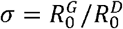.

We formulate the fitness inference framework under the following base case assumptions: (1) both D614 and G614 strains co-circulate locally during the period of fitness estimation; (2) non-pharmaceutical interventions (NPIs) have the same effect on the reproductive number of both strains; (3) the probability that an infected person is selected for viral sequencing is the same for both strains; (4) recovery from infection with either strain provides protection against reinfection of both strain during the period of estimation; and (5) the fitness of G614 does not depend on age, and age-specific susceptibility to infection is the same for both strains.

Under the base case assumptions, the next generation matrix (NGM) of infections by the G614 strain is *σ* times that of the D614 strain. As the pandemic unfolds, the proportion of G614 infections at time *t*, denoted by *ρ(t)*, will increase towards 1 if *σ* >1, remain at the same level if *σ* =1 and decline towards 0 if *σ* <1. In our previous work, we have shown that *ρ(t)* can be well-approximated using the equation:

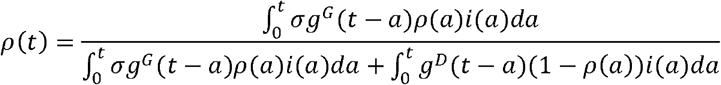

where *i(t)* is the total incidence rate (i.e. including both strains), *g*^*D*^ and *g*^*G*^ are the generation time distribution for D614 and G614 infections (assumed to be gamma distributions withas the ratio of the mean of g^*G*^ to that of *g*^*D*^), respectively. We assumed that had mean 5.4 days and standard deviation 3.8 days (estimated from empirical data ^17,32^), and *g*^*D*^ and g^*G*^ had the same coefficient of variation.

To assess the effects of importations and introductions dominated by G614 since late February for most countries in Europe and the US, we modified the equation for *ρ*(*t*) to include an imported force of infection by G614, which was *φ*_*G*_ times of the local incidence rate:

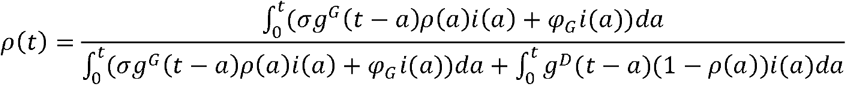

㏆G was then estimated with other parameters in the inference with the likelihood specified below.

### Inference of the G614 fitness in transmission

Our method requires two streams of data. The first data stream is the incidence rate *ĩ (t)* or its proxy, e.g. using the daily number of COVID-19 confirmed cases or deconvoluting the daily number of COVID-19 deaths with the time between infection and confirmation or death. We denote this data stream by 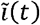 In the base case analysis, we obtained time series of COVID-19 confirmed deaths from situation updates published by World Health Organization as the proxies. We assume that the distribution of the time between infection and death is gamma with mean and standard deviation (SD) of 28 and 8.4 days (Figure 2). We use probability density function of the time from infection to deaths to deconvolute the time series of the daily number of deaths to reconstruct an epidemic curve of daily number of new infections ^33^. We only used the time series of COVID-19 confirmed cases in the sensitivity analysis because it is more often confounded with temporal fluctuations in reporting rate and testing capacity ^25^, but our previous simulations had shown that our method is robust against these fluctuations ^16^.

The second stream is the detections of D614G mutation where 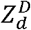 and 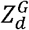 are the number of SARS-CoV-2 isolates among phylogenetic reconstructed clusters sampled on day with D614 and G614 respectively (Figure 2). Please refer to the previous section “Phylogenetic reconstruction of D614 and G614 clusters” for details of cluster reconstruction. We selected data from ten countries which had cocirculation of D614 and G614, namely Australia, Belgium, Denmark, Iceland, India, Netherlands, Spain, Portugal, the UK and the US.

In the base case, we assumed there is 7 days of delay between infection and sample collection for sequencing in the base case analysis. Similarly, we assumed on average the time between infection and reporting is 7 days and the time between infection and death is 28 days. We performed a sensitivity analysis of the time between infection and these key events: 1) we assumed the time between infection and sampling is 5, 7, 9 and 12 days with coefficient of variation of 0.3; 2) we assumed the time between infection and reporting is 5, 7, 9 and 12 days with coefficient of variation of 0.3; 3) we assumed the time between infection and death is 21, 28, and 35 days with coefficient of variation of 0.3. The estimation of G614 fitness is not sensitive to the assumptions about the time between infection and these key events.

We did not include China and other East Asian countries in the analysis because no continuous cocirculation were detected in most Asian countries and there is not enough information from GISAID to avoid misclassifying sequences from imported cases as those from local cases. We substitute *i(t)* with 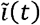 and denote the approximation of *ρ(t)* by 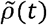. The approximate likelihood is

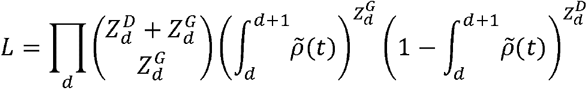

With this likelihood, the inference was performed in a Bayesian framework with non-informative priors using Markov Chain Monte Carlo.

### Data sharing statement

We collated all data from publicly available data sources. All the information that we used is available in the main text or the supplementary materials.

## Contributors

TTYL, KL, JTW and GML designed the experiments. KL, YP and TTYL collected data and performed sequence alignment and phylogenetic analysis. KL and JTW analyzed epidemiological data. KL, JTW, TTYL, and GML interpreted the results and wrote the manuscript.

## Supplementary information

### The infection fatality risks in locations predominated by D614 or G614

To test the hypothesis that the G614 mutation might affect the clinical severity of SARS-CoV-2 infection, we estimated the infection fatality risks (IFRs) as the ratio of laboratory-confirmed deaths to the estimated number of infections in locations or settings with COVID-19 circulation predominated by D614 or G614. The daily number of confirmed deaths were obtained from the websites of local public health agencies. For locations where extensive contact tracing and testing had been conducted, the number of infections were estimated as the number of reported infections; for locations where seroprevalence studies had been conducted, the number of infections were estimated by the product of the seroprevalence and the population size accordingly. We assumed on average it takes 21 days for infected individuals to develop consistently detectable antibodies after infection ^34^, and the time between infection and deaths is 28 days^17^.

Among locations where COVID-19 circulation was dominated by D614, we estimated that IFR ranged from 0.90% (0.75-1.06) in mainland Chinese provinces outside Hubei to 1.0% (0.40-2.04) among passengers from the Princess Diamond Cruise (Table 2). The IFR estimates were lower but not significantly different among locations where COVID-19 circulation was dominated by G614, ranging from 0.43% (0.37-0.56) in Geneva, Switzerland to 0.83% (0.65-1.10) in New York City, US (Table S2).

**Figure S1.**
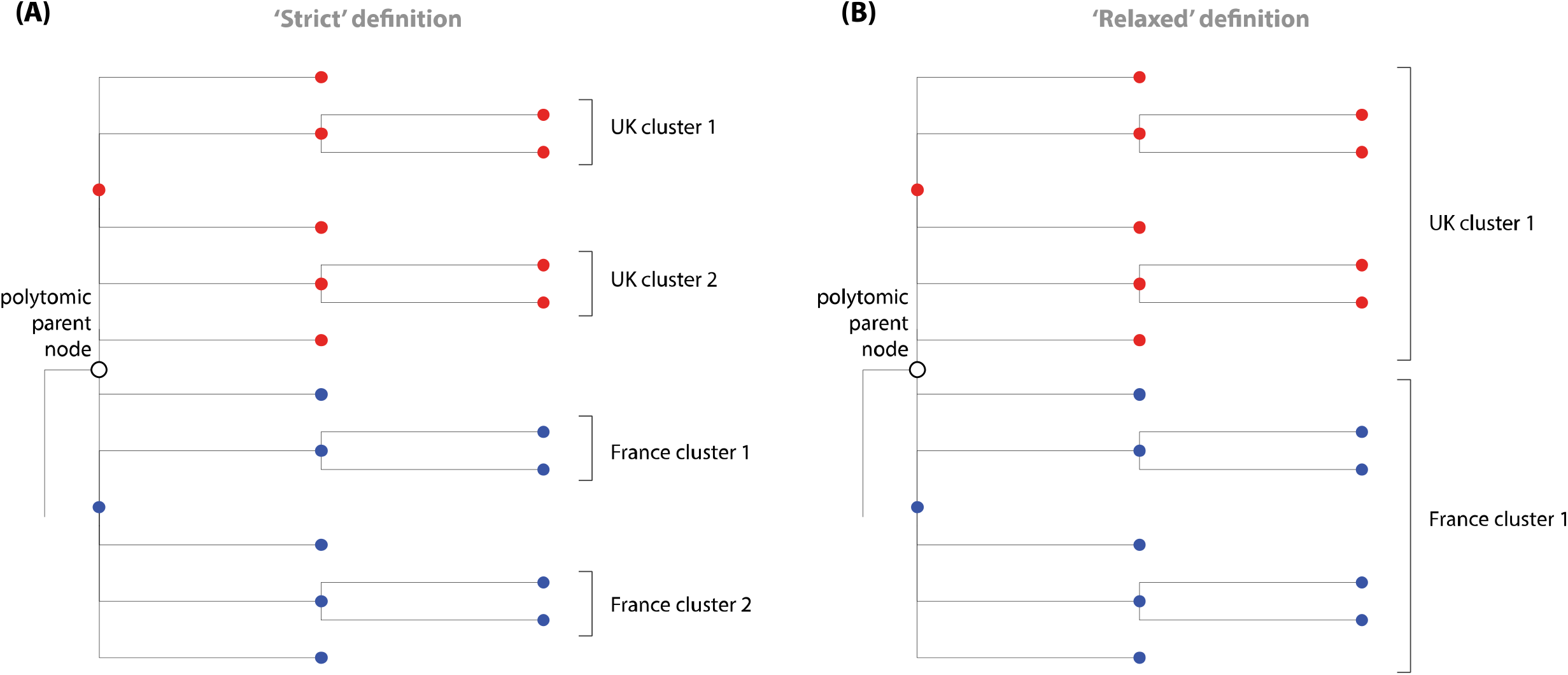
Illustration of ‘strict’ and ‘relaxed’ definitions of transmission clusters reconstructed with phylogenetic methods. Solid circles in red and blue colors are sequences from UK and France respectively. In ‘strict’ definition, only multiple sequences from the same country sharing strictly monophyletic relationship are considered as a transmission cluster. In ‘relaxed’ definition, cluster and non-cluster sequences of the same country are aggregated into a larger cluster if they share the same parent node (e.g. the open circle in the tree) even if it is a polytomy and consists of child nodes from different countries.

**Figure S2.**
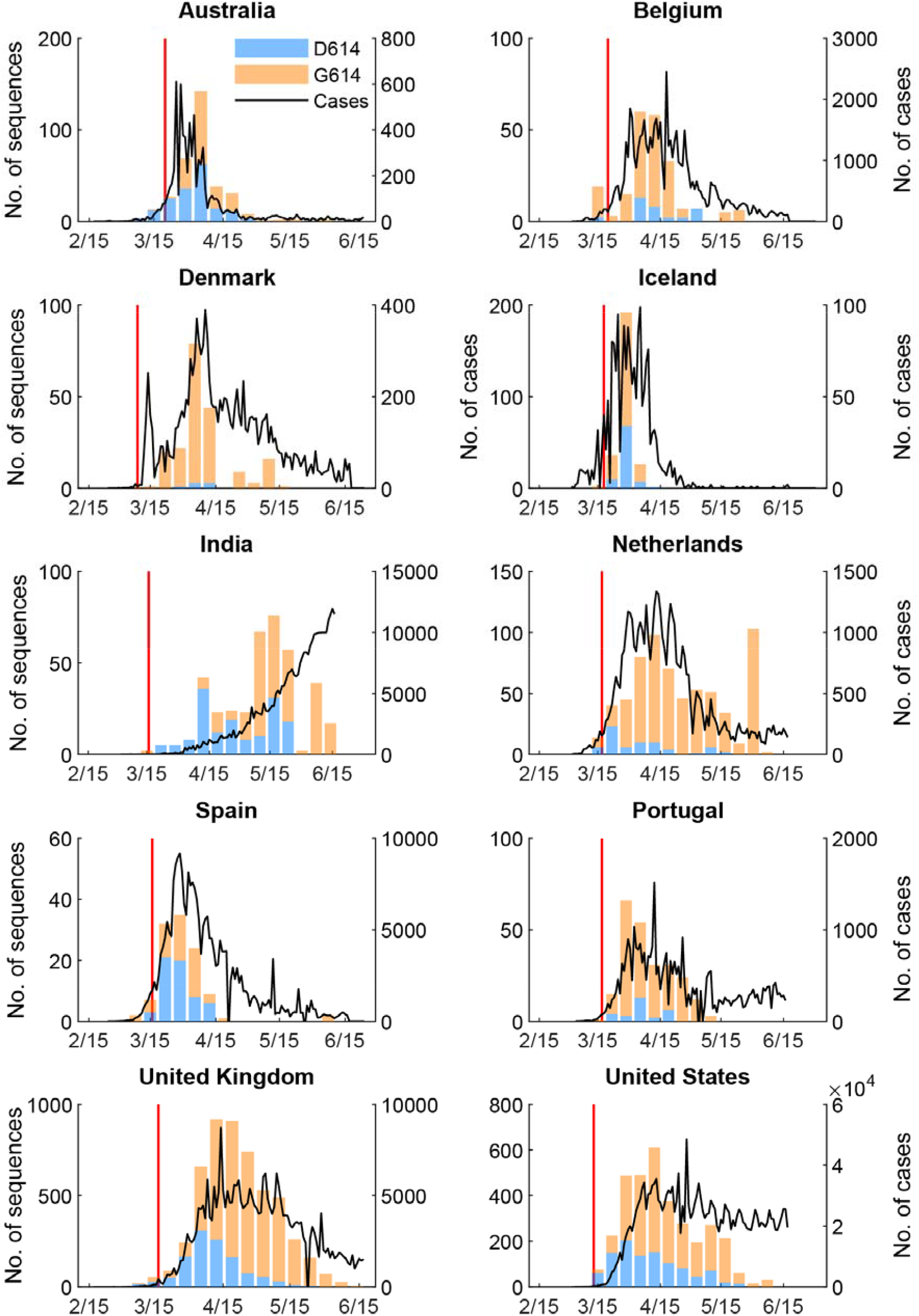
Weekly number of confirmed COVID-19 cases and the weekly number of D614 and G614 sequences from clusters with two or more cases, submitted by Australia, Belgium, Denmark, Iceland, India, Netherlands, Spain, Portugal, UK and US. Clusters with two or more sequences on GISAID were defined by phylogenetic methods with “strict” criteria. Each cluster stems from one or a small number of introductions and at least one transmission chain can be reconstructed from sequences within the same cluster. Only clusters sampled during the co-circulating period of D614 and G614 strains in each country were included in the analysis. Only countries with more than 100 sequences from at least 5 co-circulating clusters were included in the analysis. The red lines indicated the date when major travel restriction from or to countries of European Union started.

**Figure S3.**
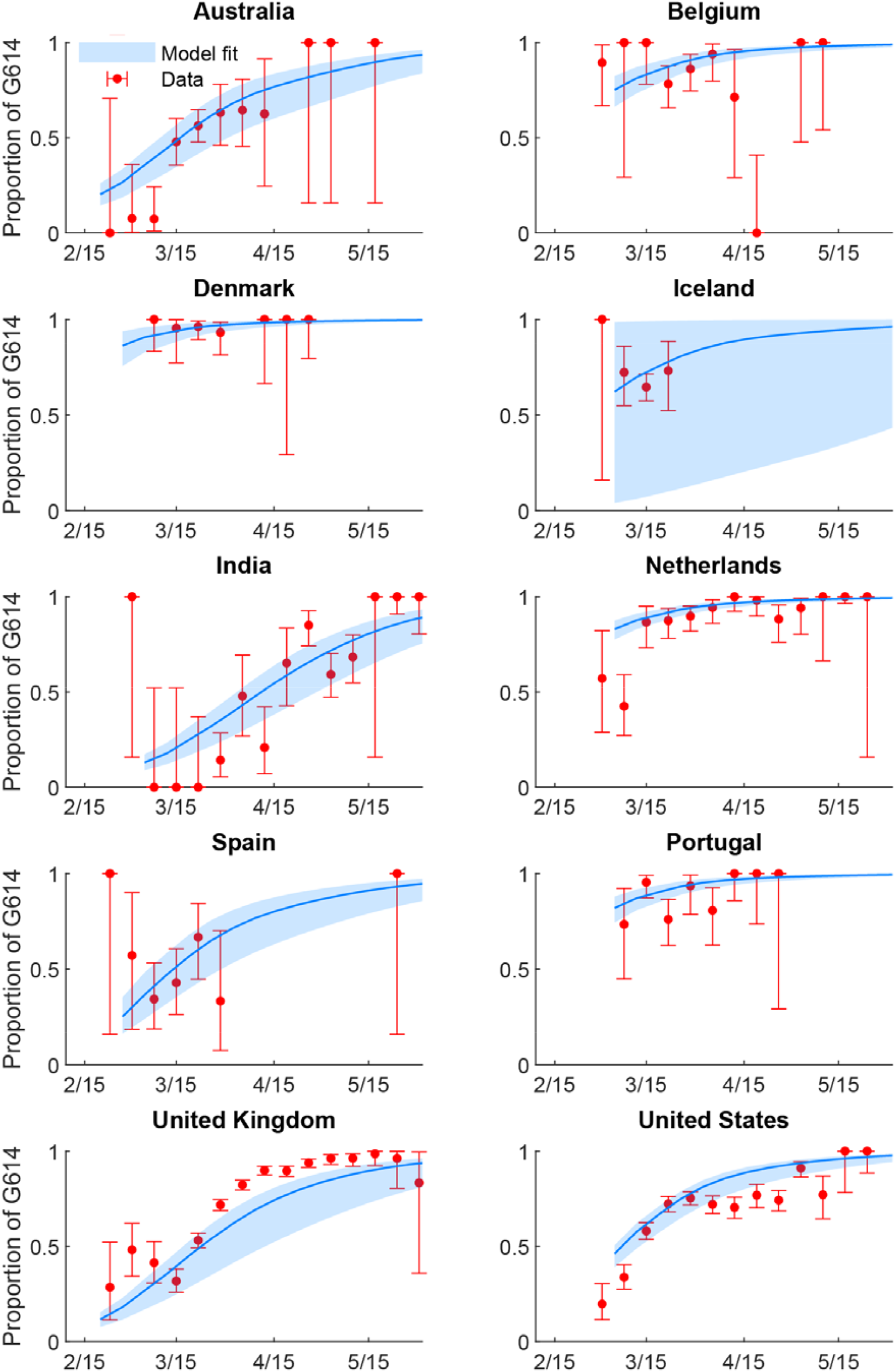
The weekly proportion of G614 infections between late January and early May when both D614 and G614 strains cocirculated. The time series of confirmed COVID-19 cases was used in the estimation. The red circles and error bars indicated the observed proportion with 95% binomial CIs among sequence data. The blue lines and shades indicated the posterior mean and 95% CrI of the estimates.

**Figure S4.**
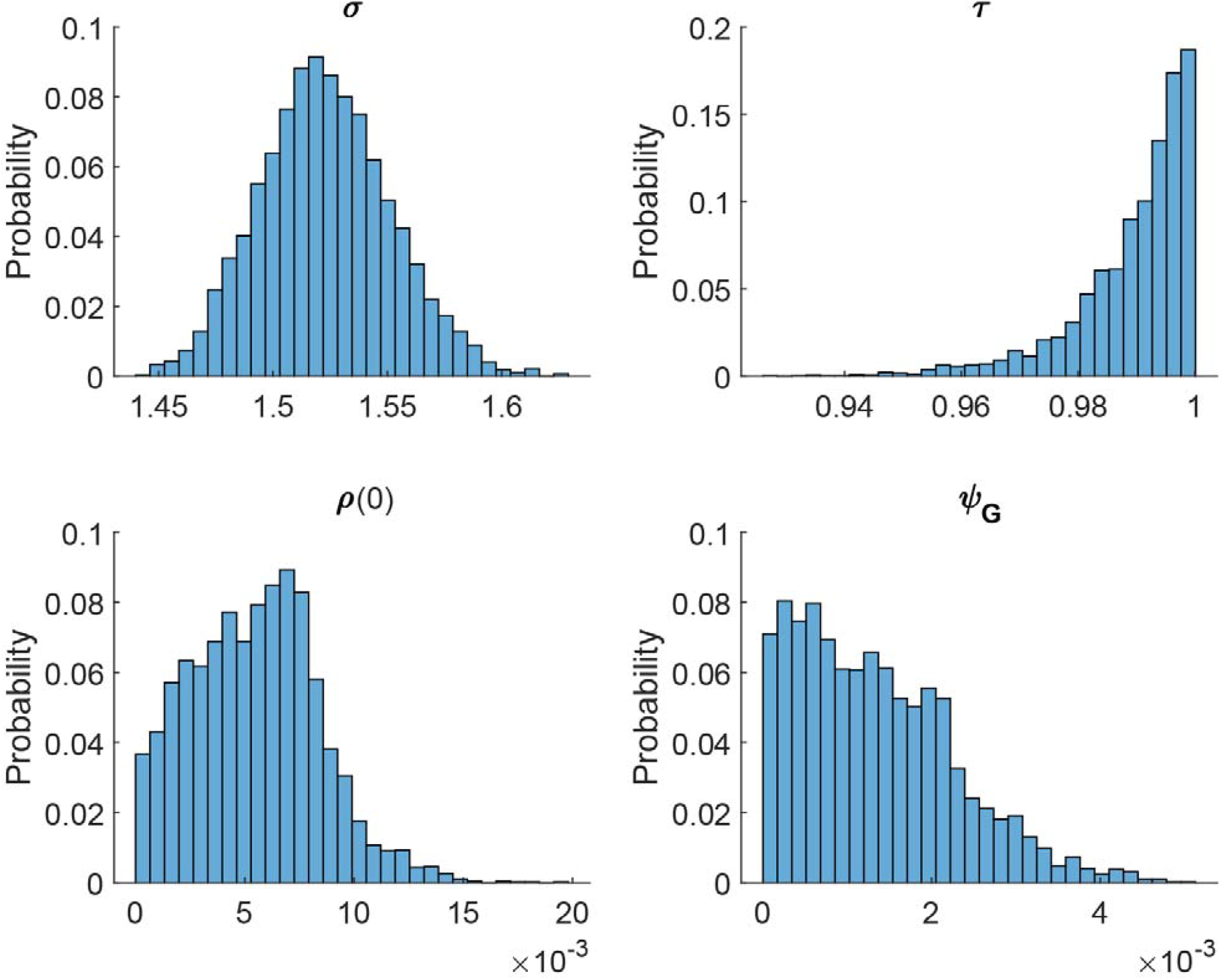
The posterior distribution of parameters if the force of infection of G614 importations is included in the fitness estimation in the UK. The base case includes clusters with ≥2 sequences in each cluster using the “strict” cluster definition assuming index case in each cluster was included. Clusters were reconstructed from sequences sampled from England, Scotland, Wales and Northern Ireland.

**Table S1.** **The posterior distribution of parameters if the force of infection of G614 importations is included in the fitness estimation in the ten selected countries**

| Parameters | Country | Posterior mean (95% CrI) |
| --- | --- | --- |
| The ratio of the basic reproduction number of the G614 strain to that of the D614 strain $\sigma$ | | 1.273 (1.233-1.313) |
| The ratio of the mean generation time of the G614 strain to that of the D614 strain $\tau$ | | 0.993 (0.961-1.000) |
| The proportion of G614 infections when both D614 and G614 started to cocirculate $\rho(0)^*$ | Australia | 0.099 (0.063-0.145) |
|  | Belgium | 0.627 (0.529-0.717) |
|  | Denmark | 0.841 (0.724-0.926) |
|  | Iceland | 0.497 (0.022-0.978) |
|  | India | 0.023 (0.002-0.068) |
|  | Netherlands | 0.745 (0.675-0.810) |
|  | Spain | 0.173 (0.106-0.253) |
|  | Portugal | 0.745 (0.656-0.821) |
|  | United Kingdom | 0.021 (0.001-0.068) |
|  | United States | 0.386 (0.351-0.417) |
| The scaling factor of the force of infections of G614 importations $\varphi_G$ | | 0.017 (0.003-0.027) |

\* Sampling dates of the 1<sup>st</sup> sequence in D614 and G614 cocirculating clusters included in the analysis of each country are the same as Table 1.

**Table S2.** **Estimated infection fatality risks in countries or regions with COVID-19 circulation predominated by D614 or G614**

| Country/Region/Setting | Circulation period of interest | Strain | No. of confirmed deaths | No. of infections or seroprevalence (95% CI) | IFR (95% CI) | Method/Source |
| --- | --- | --- | --- | --- | --- | --- |
| Wuhan, Hubei Province, China | Jan-Mar 2020 | D614 | 3,869 | To et al<br>3.76% (2.21-5.95)<br>Xu et al<br>3.67% (2.71-4.86) | 0.93% (0.59-1.58)<br>0.95% (0.72-1.29) | <sup>35-37</sup> |
| Provinces outside Hubei in mainland China | Jan-Mar 2020 | D614 | 135 | 15,076* | 0.90% (0.75-1.06) | <sup>17</sup> |
| Diamond Princess Cruise | Feb 2020 | D614 | 7 | 705 | 0.99% (0.40-2.04) | <sup>38</sup> |
| Washington State, US | Feb-Apr 2020 | D614 & G614 | 610<br>(as of 8 Apr §) | 1.13% (0.70-1.94) | 0.71% (0.41-1.14) | <sup>39</sup> |
| New York City, New York State, US | Feb-Apr 2020 | G614 | 13,495<br>(as of 26 Apr §) | 19.3% (14.6-24.9) | 0.83% (0.65-1.10) | <sup>40</sup> |
| Geneva, Switzerland | Feb-May 2020 | G614 | 230<br>(as of 16 May §) | 10.8% (8.2-12.3) | 0.43% (0.37-0.56) | <sup>41</sup> |
| London, UK | Feb-May 2020 | D614 & G614 | 7,957<br>(as of 15 May §) | 17.5% (13.4-22.8) | 0.51% (0.39-0.67) | <sup>42</sup> |

\* Assuming the number of reported local cases was close to the number of local infections because the ascertainment rates were ~100% in provinces outside Hubei given the intensive and proactive case finding
§ 7 days after the last week of the last estimate of seroprevalence, assuming it takes 21 days on average to develop consistently detectable antibodies after infection and the time between infection and deaths is 28 days

